# Decomposed interaction testing improves detection of genetic modifiers of the relationship of dietary omega-3 fatty acid intake and its plasma biomarkers with hsCRP in the UK Biobank

**DOI:** 10.1101/2024.09.09.24313018

**Authors:** Kenneth E. Westerman, Chirag J. Patel, James B. Meigs, Daniel I. Chasman, Alisa K. Manning

## Abstract

Discovery and translation of gene-environment interactions (GxEs) influencing clinical outcomes is limited by low statistical power and poor mechanistic understanding. Molecular omics data may help address these limitations, but their incorporation into GxE testing requires principled analytic approaches. We focused on genetic modification of the established mechanistic link between dietary long-chain omega-3 fatty acid (dN3FA) intake, plasma N3FA (pN3FA), and chronic inflammation as measured by high sensitivity CRP (hsCRP). We considered an approach that decomposes the overall genetic effect modification into components upstream and downstream of a molecular mediator to increase the potential to discover gene-N3FA interactions. Simulations demonstrated improved power of the upstream and downstream tests compared to the standard approach when the molecular mediator for many biologically plausible scenarios. The approach was applied in the UK Biobank (N = 188,700) with regression models that used measures of dN3FA (based on fish and fish oil intake), pN3FA (% of total fatty acids measured by nuclear magnetic resonance), and hsCRP. Mediation analysis showed that pN3FA fully mediated the dN3FA-hsCRP main effect relationship. Next, we separately tested modification of the dN3FA-hsCRP (“standard”), dN3FA-pN3FA (“upstream”), and pN3FA-hsCRP (“downstream”) associations. The known *FADS1-3* locus variant rs174535 reached *p* = 1.6x10^-12^ in the upstream discovery analysis, with no signal in the downstream analysis (*p* = 0.94). It would not have been prioritized based on a naïve analysis with dN3FA exposure and hsCRP outcome (*p* = 0.097), indicating the value of the decomposition approach. Gene-level enrichment testing of the genome-wide results further prioritized two genes from the downstream analysis, *CBLL1* and *MICA*, with links to immune cell counts and function. In summary, a molecular mediator-focused interaction testing approach enhanced statistical power to identify GxEs while homing in on relevant sub-components of the dN3FA-hsCRP pathway.

## Background

Gene-environment interactions (GxEs), in which the relationship between an exposure and outcome is modified by a genetic variant, continue to hold promise for the development of more precise clinical and behavioral interventions for the prevention of cardiometabolic disease. Compared to intervention trials, observational datasets allow for the exploration of GxEs in much larger sample sizes and with a broader array of exposures and thus a greater opportunity to uncover new biology. However, these analyses are still limited in statistical power (Gauderman et al., 2017).

Molecular omics data are being increasingly collected in observational cohort studies, with promise to add mechanistic insight and dynamic longitudinal measures to genotype-only analyses. This potential applies directly to some of the challenges present in GxE studies. Molecular data can increase power for detection by acting as objective proxies for exposures (e.g., lifestyle or environment) that otherwise require noisy estimates based on self-report (Patel et al., 2017). They also represent intermediates on biological pathways linking genotypes and exposures to outcomes of interest, allowing for mechanistically informative analyses (Mohammadi-Shemirani et al., 2023). However, such molecular data require associated analytical approaches to coherently incorporate them into GxE testing.

A particularly straightforward example of such a mediated pathway is the relationship between dietary long-chain omega-3 fatty acid (dN3FA) intake, plasma N3FA (pN3FA), and chronic inflammation (Calder, 2015). dN3FA intake, coming primarily from oily fish and fish oil supplements, is a primary determinant of pN3FA, which acts as a physiological N3FA status indicator and is sometimes used as a dietary intake proxy. pN3FA is consistently associated with lower hsCRP, and long-chain N3FA supplementation has been shown to decrease the chronic inflammatory biomarker high-sensitivity C-reactive protein (hsCRP), though this finding is less consistent. Genetic variation, especially at the fatty acid desaturase (*FADS*) locus, may play a role in this variable response to dN3FA (Schulze et al., 2020).

We reasoned that a decomposed interaction testing approach, separating an exposure-outcome pathway into components upstream and downstream of a molecular mediator, would increase the potential to discover associated gene-N3FA interactions impacting hsCRP. We first use simulations to illustrate the expected gain in statistical power to detect interactions using this approach. We proceed to explore genetic modification of the dN3FA-hsCRP relationship after its decomposition into dN3FA-pN3FA (upstream) and pN3FA-hsCRP (downstream) sub-pathways, uncovering interactions at variants and genes that would not have been found using a standard GxE testing approach.

## Methods

### Simulations

Simulations were performed to understand how statistical power for interaction detection changes when varying a small set of underlying parameters governing the strength of the interactions, mediating pathways, and measurement error. First, genotypes (G) were generated from a binomial distribution with a minor allele frequency of 0.25 and exposures (E) were generated from a standard normal distribution. Next, a mediator (M) was generated, incorporating signal from a G*E product term (for upstream simulations) or from an E main effect (for downstream simulations). From this, a “measured” mediator (M_measured_) was generated, tracking the true M but with added noise according to a specified intraclass correlation coefficient (ICC_M_). Finally, an outcome (Y) was generated, incorporating signal from a main effect of M (for upstream simulations) or from a G*M product term (for downstream simulations). For each scenario, interaction terms were tested for significance in two of three regressions: Y∼G + E + GE (standard), M_measured_ ∼G +E + GE (upstream only), and Y∼G + M + GM_measured_ (downstream only). Finally, power for each scenario was calculated as the fraction of tests passing the chosen significance threshold out of the total number of repeated simulations.

For simplicity, in the primary set of simulations, the sample size was fixed at N = 1,000, the significance threshold was set to 0.05, and the number of repeated simulations per scenario was fixed at 500. For upstream simulations, three parameters were varied: the proportion of variance in M explained by the G×E interaction, the proportion of variance in Y explained by M, and the measurement error in M (ICC_M_). For downstream simulations, three parameters were varied: the proportion of variance in M explained by E, the proportion of variance in Y explained by the G×M interaction, and the measurement error in M (ICC_M_). We note that, despite the small sample size used here compared to biobank datasets, the relative changes in power based on simulated interaction strength, degree of pathway mediation, and measurement error should be consistent.

### UK Biobank population and genotype data

The primary analysis was conducted under a Not Human Subjects Research determination for UKB data analysis (NHSR-4298 at the Broad Institute of MIT and Harvard) and UK Biobank application 27892. UKB is a large prospective cohort with both deep phenotyping and molecular data, including genome-wide genotyping, on over 500,000 individuals ages 40-69 living throughout the UK between 2006-2010 (Sudlow et al., 2015). Genotyping, imputation, and initial quality control on the genetic dataset have been described previously (Bycroft et al., 2018). We used a multi-ancestry sample of individuals that had not withdrawn consent by the time of analysis. Additionally, we subset to a group of unrelated samples (by including only those that were used for genetic principal components analysis during central genetic data preprocessing) and removed participants who were pregnant or had diabetes, coronary heart disease, liver cirrhosis, or cancer. Genetic variants with minor allele frequency (MAF) >1% in the full analysis population were included in genome-wide and follow-up studies.

### Phenotypic data

The primary outcome trait, hsCRP, was originally measured in plasma using an immunoturbidimetric assay (Beckman Coulter AU5800). Values were log-transformed and outliers (more than 5 standard deviations from the mean of the log-transformed values) were removed prior to analysis.

N3FA intake data came from multiple self-reported sources. All participants completed a 30-item food frequency questionnaire (FFQ) at the baseline assessment center visit, which has been validated for reliably ranking participants according to intake of major food groups but is not sufficient for calculating specific nutrient intake estimates (Bradbury et al., 2018). Estimates for typical intake of both oily fish and non-oily fish (servings/day; fields 1329 and 1339) were retrieved from this FFQ. Fish oil supplementation was recorded based on reported use as a medication, with variables derived from both touchscreen questionnaire (UKB field 6179) and verbal interview (field 20003).

Additional covariates collected for analysis included genetically-determined sex, age, age^2^, a sex-by-age product term, income (5 categories), educational attainment (6 categories), smoking (categorical: never, past, or current), alcohol intake (categorical: weekly frequency estimates), and additional diet variables from the FFQ (cooked vegetables, raw vegetables, fresh fruit, processed meat), and a categorical diet variable (bread type; wholemeal or wholegrain bread versus other types). For variables coded as categorical, ambiguous categories such as “do not know” or “prefer not to answer” were left as non-missing to allow them to constitute an independent category for adjustment.

### Plasma N3FA measurements

Various measurements of plasma N3FA status were available from the Nightingale platform (N = 199,059), These data were preprocessed using the *ukbnmr* R package (Ritchie et al., 2023), which includes imputation of zero values, log-transformation, adjustment for key batch variables such as shipment plate and time between sample preparation and measurement, and transformation back into absolute concentrations. Ultimately, relevant available N3FA species included: direct N3FA concentrations, docosahexanoic acid (DHA), and both of these quantities as fractions of total fatty acids and total polyunsaturated fatty acids. Non-DHA pN3FA was calculated by subtracting DHA% from total N3FA%. Eicosapentanoic acid was not measured directly but should constitute approximately two thirds of this non-DHA quantity (Abdelmagid et al., 2015).

An overall estimate of dietary N3FA intake (“dN3FA”) was calculated as a weighted sum of four key sources: oily and non-oily fish intake based on FFQ, touchscreen-reported fish oil intake, and verbal interview-reported fish oil intake. To determine the relative contribution of each source, total N3FA% was regressed on the four dietary sources in the full population. The dN3FA variable was then calculated as a linear combination of these components, weighted by their corresponding multivariable regression effect estimates. Missing values for fish oil variables were imputed as “no intake” when taking this weighted sum.

### Main effect analysis and mediation testing

All statistical analyses were performed using R version 4.2.2 (R Core Team, 2022) unless otherwise noted. Linear regression models were used to understand the associations between various self-reported dietary N3FA sources and hsCRP using standard model-based standard errors (in contrast to the robust standard errors used in genome-wide interaction testing) and the covariates described above. Supp. Fig. S2 shows the directed acyclic graph and preliminary main effect regression results used to guide the choice of covariates. Initial models included additional adjustment for assessment center variable (one indicator variable per center), but this adjustment was removed from downstream analysis due to the minimal impact on regression estimates.

Mediation analysis examined the degree to which the plasma fatty acids mentioned above mediated the relationship between dN3FA and hsCRP. Mediation tests were performed using the *mediation* package for R (Imai et al., 2010; Tingley et al., 2014), using robust standard errors and 20 Monte Carlo draws. Exposures and outcomes were scaled to mean zero and standard deviation one to create comparable mediation effect estimates across exposures. The output of each analysis included estimates of the total effect, the average causal mediation effect (ACME), and the direct effect. We note that, given the continuous exposure and outcome variables explored here, this mediation framework reduces conceptually to the linear structural equation modeling approach originally described by Baron and Kenny (Baron & Kenny, 1986).

### Gene-environment interaction modeling

Genome-wide interaction studies (GWIS) were performed using GEM v1.5.2 (Westerman et al., 2021) with robust standard errors. The primary interaction model was as follows:

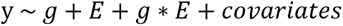

where y represents the outcome of interest, g represents the imputed genotype dosage, and E represents the exposure of interest. Covariates were the same as used for main effect models, with the addition of 10 genetic principal components as calculated centrally by the UKB team. GWIS were performed for three pathways: “standard” (E = dN3FA, Y = hsCRP), “upstream” (E = dN3FA, Y = pN3FA), and “downstream” (E = pN3FA, Y = hsCRP). Significance was assessed based on a standard genome-wide threshold of *p* < 5×10^-8^. GWIS results were pruned using PLINK 2.0 (Chang et al., 2015), using an LD reference panel consisting of a random 20,000-participant subset of the UKB and with parameters as follows: index variant *p*-value threshold = 5×10^-8^, LD r^2^ threshold = 0.2, and clumping radius = 5,000 kb. Sensitivity models at top loci included adjustment for: (1) exposure-by-gPC interaction terms, (2) genetic interaction terms for all covariates, and (3) body mass index (BMI) as a measure of adiposity.

GWIS results were subject to enrichment analysis to prioritize genes with enrichment of interaction signal in the surrounding genetic region. Interaction *p*-values from the GWIS were used as input to the MAGMA program (de Leeuw et al., 2015), using the same LD reference panel as used for pruning and gene regions defined from 2kb upstream to 1kb downstream of the gene limits based on the NCBI database (GRCh37). Sensitivity analyses as described above were performed using the most significant variant from each gene-based finding.

## Results

Simulations revealed the extent to which the decomposed approach is advantageous over the standard GxE testing approach as a function of the magnitude of the genetic interaction and the relationships between the exposure (E), mediator (M), and outcome (Y). As shown in Fig. 1, upstream interaction tests were more powerful when the true genetic interaction was with E (rather than M). Intuitively, the power of the upstream test matched that of the standard test as the M-Y relationship became stronger (i.e., in the limit that M fully determines Y). However, at more plausible values, the difference in power was substantial: when the GxE explained 0.5% of the variance in M, which then explained only 10% of the variance in Y, the upstream test was more powerful by a factor of 1.79 (38% versus 21% power). Even more stark patterns were observed for the downstream tests as a function of the strength of the E-M relationship. Power for the downstream test was equal when E fully determined M, but was greater by a factor of 4.72 (43% versus 9% power) when we simulated a GxM explaining 0.5% of the variance in Y and an E explaining 10% of the variance in M. We note that the simulations presented in Fig. 1 assume perfect measurement of the mediator; power of the decomposed approach decreases as this measurement error increases (see Supp. Fig. S1). Nonetheless, these results support the greater power of the decomposed approach for GxE discovery in most scenarios in which molecular mediators are known and measured.

**Figure 1:**
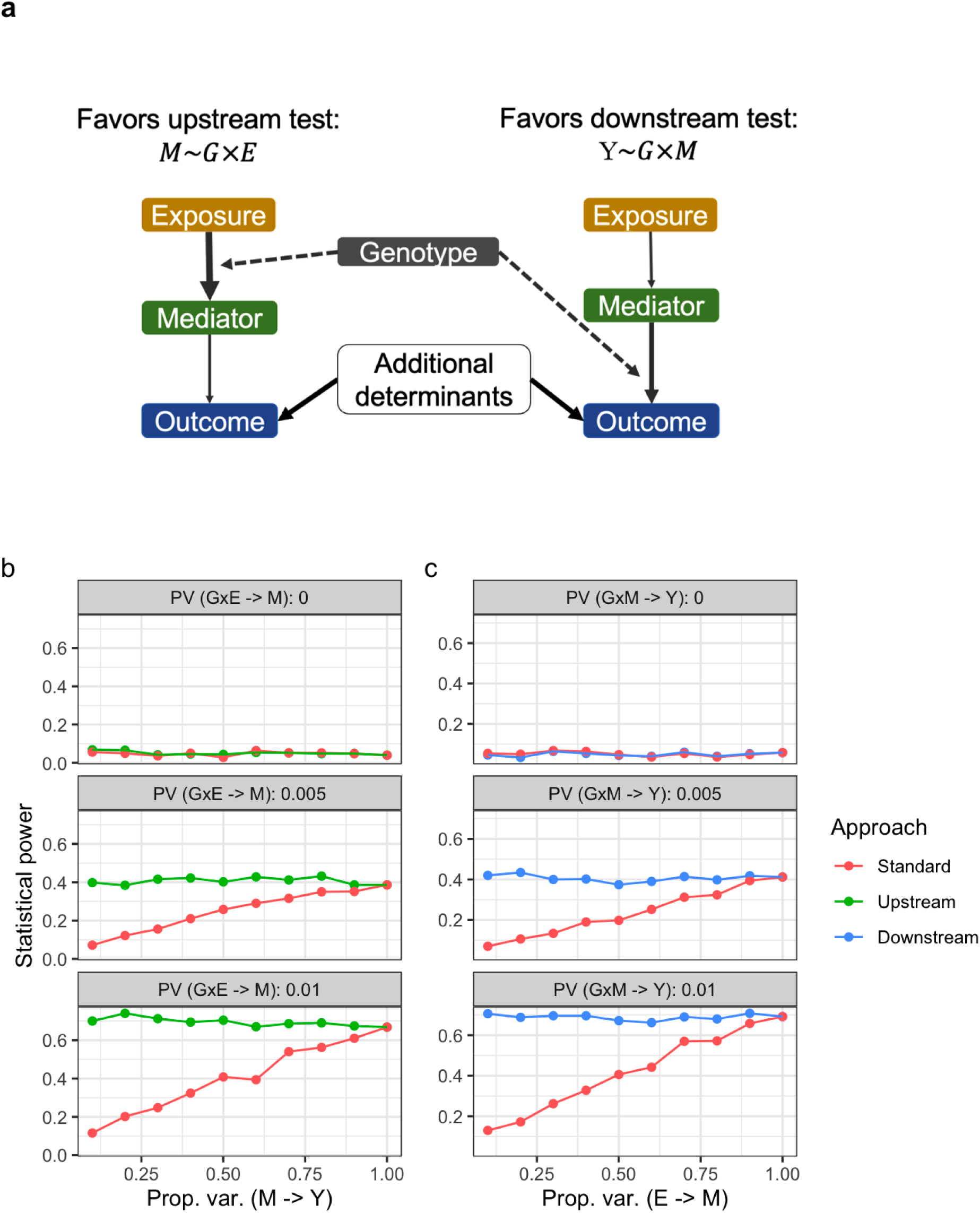
Power simulations and conceptual basis for the decomposed GxE testing approach. a) Conceptual diagram indicates the relationships between the exposure (E), mediator (M), and continuous outcome (Y), with line thickness denoting proportion of variance explained. Dotted lines correspond to interaction effects. b,c) Power plots display simulation results. *X*-axes correspond to the strength (quantified by proportion of variance explained) of the M-Y relationship (upstream, left) or E-M relationship (downstream, right). Faceted panels correspond to the strength of the simulated genetic interaction with E (upstream) or M (downstream). No measurement error is included in the simulations summarized here (see Supp. Fig. S1).

A summary of the multi-ancestry UKB population can be found in Supp. Table S1. Preliminary regressions confirmed the expected negative association between dietary N3FA sources (fish and fish oil) and hsCRP, with the association partially attenuated by adjustment for confounders (Supp. Fig. S2). Of the available pN3FA measures, total N3FA, as a percentage of total plasma fatty acids, showed the strongest correlations with dietary N3FA sources. Based on this, an overall estimate of dietary N3FA intake (“dN3FA”) was calculated as a weighted sum of four key sources (oily and non-oily fish, touchscreen-reported fish oil intake, and verbal interview-reported fish oil intake), with oily fish contributing most substantially to the derived dN3FA estimate (Supp. Fig. S3, Supp. Table S2).

This derived dN3FA metric was positively associated with pN3FA, while both dN3FA and pN3FA were negatively associated with hsCRP (Fig. 2a-c). Mediation analysis showed that the dN3FA-hsCRP relationship was mediated by pN3FA (mediated effect: -0.036, total effect: -0.023; Fig. 2d, Supp. Table S3). This result, with a mediated effect greater than the total estimated effect, is a case of inconsistent mediation that can indicate residual negative confounding in the opposite direction of the mediated effect (MacKinnon et al., 2000). Thus, we explored several additional mediation analyses to confirm the result. As shown in Supp. Fig. S4, the total and mediated effects were larger for oily compared to non-oily fish, as expected due to the higher N3FA content. Additionally, raw vegetable intake, which is related similarly correlated with other confounding variables and outcomes (Supp. Fig. S4a), has a comparable total effect on hsCRP, but with minimal estimated mediation by pN3FA. When comparing mediation estimates across pN3FA subspecies (DHA vs. non-DHA), most of the mediated effect was attributable to DHA (Fig. 2e).

**Figure 2:**
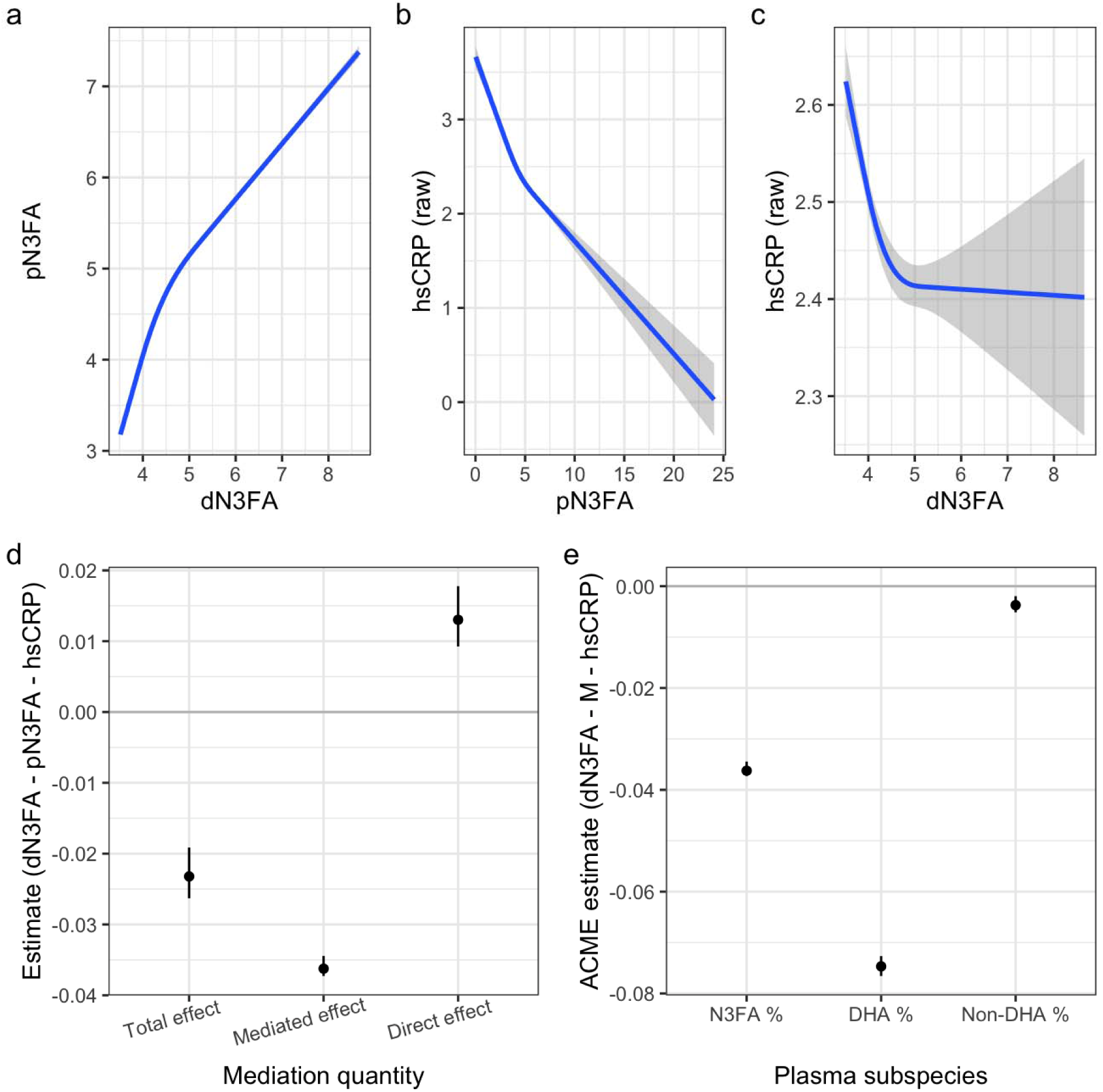
Relationships and mediation between dietary N3FA, plasma N3FA, and hsCRP. (a-c) Curves show the relationship between dN3FA and pN3FA (a), pN3FA and non-transformed hsCRP (b), and dN3FA and non-transformed hsCRP (c). Curves were estimated using a restricted cubic spline with 3 knots. *X*-axes correspond to estimated dN3FA (units of % total blood fatty acids due to the derivation of this dietary intake proxy; see Methods) or pN3FA (same units). *Y*-axes correspond to pN3FA or hsCRP (mg/L). d) Estimates and 95% CIs for the total, mediated (indirect; ACME), and direct effects from mediation analysis are shown for the pathway in which pN3FA mediates the dN3FA-hsCRP relationship. e) ACME (mediated effect) estimates are shown using either total % N3FA (i.e., total pN3FA) or subsets of this quantity (DHA and non-DHA; see Methods) as the mediating quantity.

Next, we performed a series of three GWIS, corresponding to the upstream, downstream, and standard pathways. One locus (on chromosome 11 containing the *FADS* genes) reached *p* < 5×10^-8^ in the upstream analysis, while there were none for the downstream or standard pathways (Fig. 3; Supp. Fig. S5; index variants reaching *p* < 5×10^-6^ listed in Supp. Table S4). Given the strong signal and known biology for the *FADS* locus, we explored the set of interactions at this locus more in-depth. The lead variant for the upstream analysis, rs174535, reached *p*_int_ = 1.6×10^-12^. Importantly, it showed no interaction signal for the downstream pathway (*p*_int_ = 0.94) and would not have been prioritized based on the standard pathway analysis (*p*_int_ = 0.097; Fig. 4a). This variant-specific difference in findings was supported by an additional power simulation applying relevant parameters: using a sample size of 200,000, GxE proportion of variance explained of 0.025^2^ = 0.000625, M-Y proportion of variance explained of 0.009, and MAF of 0.25, the upstream pathway had a power of approximately 1 at genome-wide significance, versus 0 for the standard pathway. Exploring the signal at this variant further, stratified plots indicated a stronger association between dN3FA and pN3FA for carriers of the pN3FA-decreasing allele (Fig. 4b). Sensitivity analyses for the interaction with rs174535 indicated robustness to additional covariates, including gPC-exposure and genotype-covariate interaction terms and adjustment for adiposity (see Methods; Supp. Table S5), and did not reveal any direct association with dietary intake behavior (from regression of dN3FA on rs174535; *p* = 0.80).

**Figure 3:**
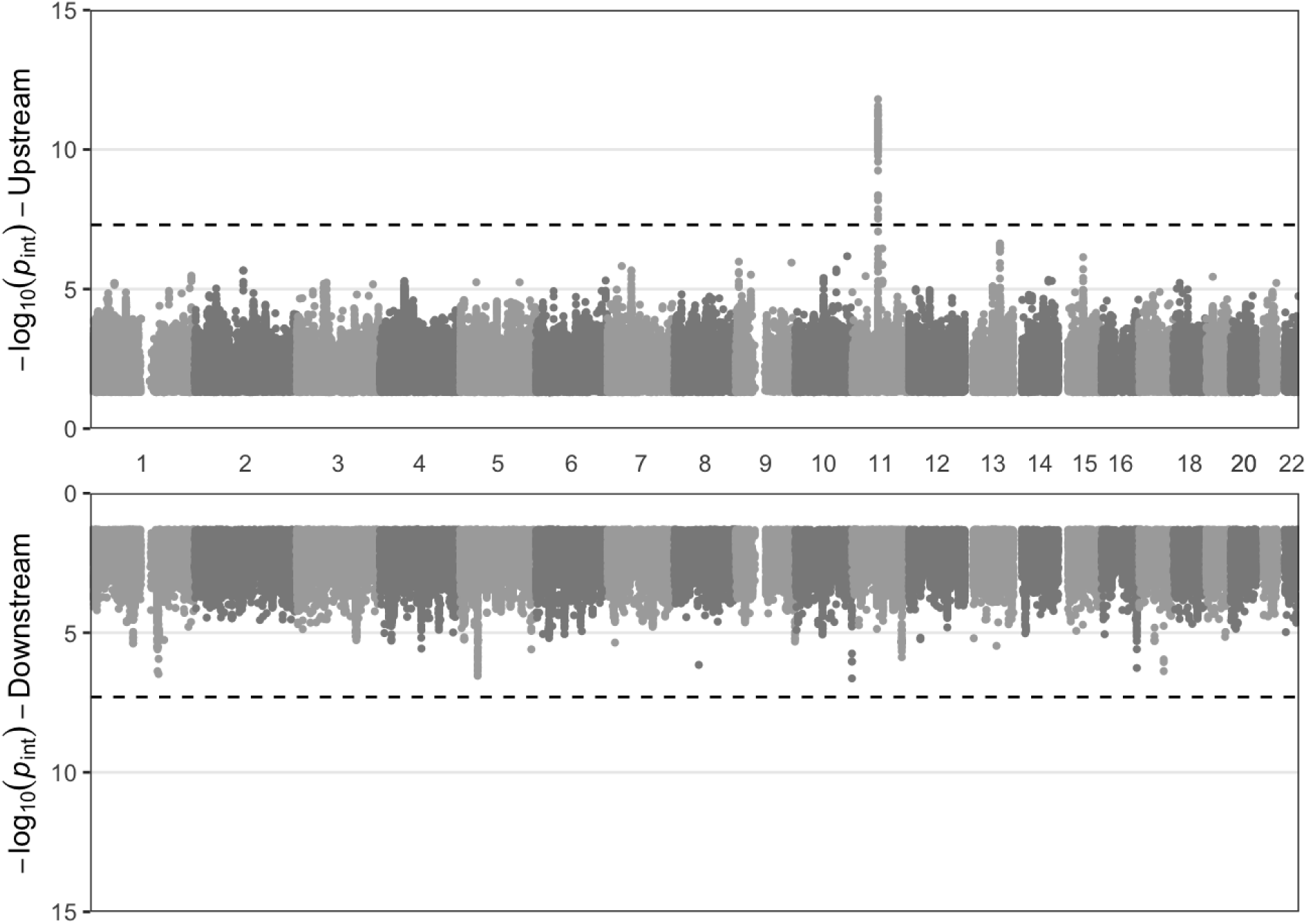
Chicago plot displays variant interaction *p*-values as a function of chromosomal position for the upstream (top) and downstream (bottom) pathways. *Y*-axes show -log(*p*) for interaction tests (based on robust standard errors), while the *x*-axis indicates chromosomal position. Dotted lines indicate the genome-wide significance threshold of 5×10^-8^. See Supp. Fig. S5 for results from the standard pathway.

**Figure 4:**
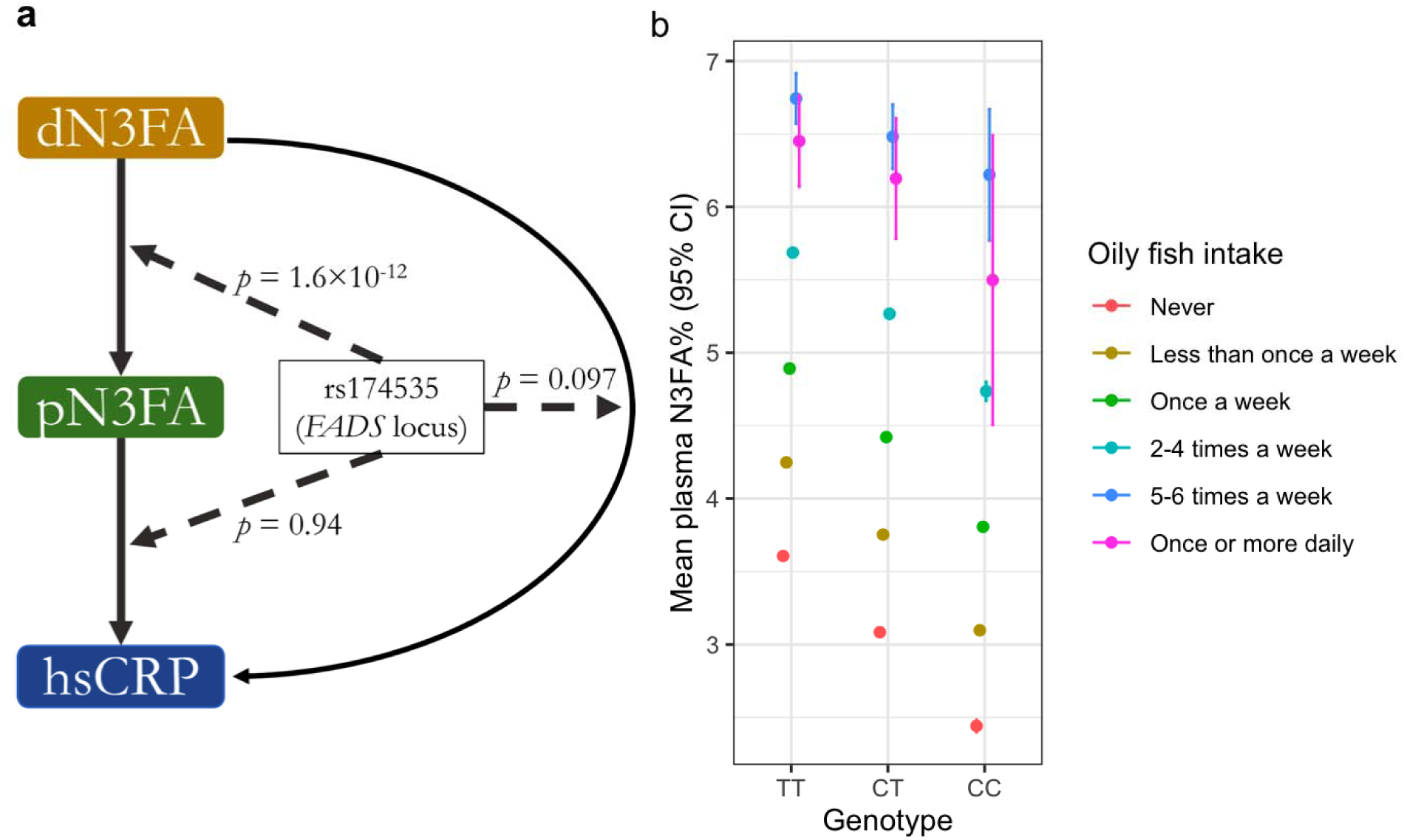
Investigation of the *FADS* locus interactions. a) Summary of the decomposed interaction testing for rs174535, including the standard, upstream, and downstream test *p*-values for interaction. b) Depiction of the interaction at rs174535 using mean pN3FA levels stratified by genotype (*x*-axis) and categories of oily fish intake (colors).

We then performed gene-level enrichment analysis using the MAGMA program to further increase power by pooling signal across variants within each gene region (Fig. 5). Using a false discovery rate threshold of *q* < 0.05 (based on the Benjamini-Hochberg method calculated separately for each of the three pathways), this analysis showed the same signals for the upstream and standard analyses (a series of genes within the larger *FADS* locus, and none, respectively). The downstream analysis revealed two additional genes: *CBLL1* (*p_gene_* = 4.9×10^-7^) and *MICA* (*p_gene_* = 3.9×10^-6^). *CBLL1* codes for a gene involved in immune-related pathways and has genetic associations with circulating immune cell counts. *MICA* also has very strong evidence for an effect on immune cell counts and codes for a gene with a key role in adaptive immune function. Thus, while having only modest genetic main effects on hsCRP itself in association studies, both genes have biologically plausible mechanisms for involvement in the modulation of inflammation by immune cells in response to changes in pN3FA status. These genes did not show any signal in gene-level enrichment of results from the standard pathway (both *p* > 0.05).

**Figure 5:**
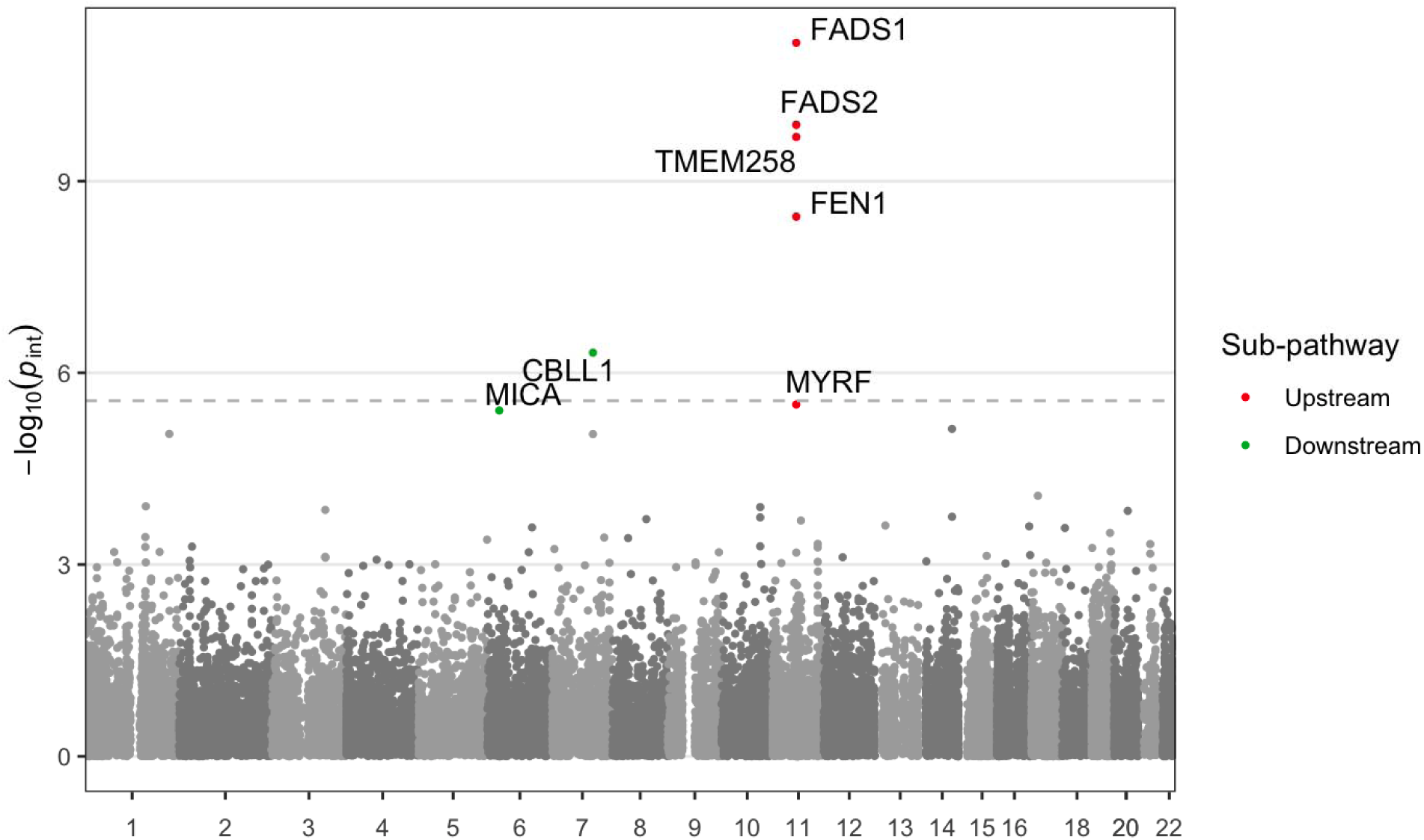
Manhattan plot displays gene-level interaction *p*-values as a function of chromosomal position for the upstream (red) and downstream (green) pathways. The *y*-axis shows -log(*p*) for gene-level enrichment tests of interaction effects from MAGMA, while the *x*-axis indicates the chromosomal position of the 5’ gene boundary. The dotted line denotes a Bonferroni significance threshold accounting for 18,249 genes tested. Colored points correspond to genes reaching FDR *q* < 0.05.

## Discussion

We explored the use of a molecular mediator-focused interaction testing approach to improve the statistical power and biological precision of GxE discovery analysis, using N3FA and hsCRP in a mechanistically informed case study. We showed that variants in the key *FADS* locus modify the dN3FA-pN3FA relationship, with an upstream interaction appearing that would not have been discovered independently of the molecular mediator. Furthermore, we described additional genes appearing in enrichment analysis of the downstream pathway analysis.

Analytical approaches combining omics and mediation analysis are increasingly used (Mohammadi-Shemirani et al., 2023), with continual methods development improving statistical models for multiple models and incomplete data (Kidd et al., 2023; Yang et al., 2021). There has been some discussion in the statistical literature of frameworks combining mediation and interaction, with distinctions made between mediation upstream or downstream of the interaction itself (Kwan & Chan, 2018; Little et al., 2012). One example from genetic epidemiology leveraged this concept to explain how sex-associated blood cell proportion differences might explain sex heterogeneity in expression-quantitative trait loci (Kasela et al., 2024). The approach described here differs in that its goal is to increase GxE discovery by leveraging knowledge about molecular mediators, rather than to explain previously observed interactions.

The strongest and only variant-specific finding was part of the upstream pathway analysis. Conceptually, this relates to the relationship between dietary intake and plasma N3FA status, which is affected by both the efficiency of absorption of N3FAs and the rate of their endogenous production from essential fatty acid precursors. Notably, this interaction signal would not have been uncovered in the standard pathway analysis (genetic modification of the dN3FA-hsCRP relationship). This lack of signal using the standard approach can be explained by the complex and multifactorial set of inputs determining hsCRP, with pN3FA explaining only a small portion of the variability in hsCRP in our dataset (0.9%). This discrepancy is supported by the *post hoc* power comparison indicating a power of 1 for the upstream pathway versus 0 for the downstream pathway. However, this does not necessarily indicate a lack of relevance of this interaction for chronic inflammation, due to both the imperfection of hsCRP as a marker of chronic inflammation and potential error in hsCRP measurement. Furthermore, by revealing modifiers of pN3FA status, this type of upstream analysis has implications for not only inflammation, but also any N3FA-related risk factor or disease state.

The *FADS* locus has one of the strongest known main effects on pN3FA (Lemaitre et al., 2011). The biological function of these genes, especially *FADS1* and *FADS2*, is related to the production of long-chain N3FAs from alpha-linolenic acid. Interactions of this locus with dN3FA intake have been explored, with inconsistent findings (Schulze et al., 2020). Based on our stratified models (such as in Fig. 4b), variants at this locus related to lower mean pN3FA also associate with a modestly stronger association between dN3FA and pN3FA, consistent with an overall endogenous feedback mechanism that enables greater absorption and incorporation of N3FA given lower baseline status. This is additionally consistent with the fact that the proteins coded for by the *FADS* genes produce long-chain N3FAs but do not act directly on absorbed dietary long-chain N3FAs. Smith and colleagues did not find evidence of a similar interaction impacting pN3FA for two variants in the *FADS1* locus in a meta-analysis of cohorts from the CHARGE consortium (Smith et al., 2015), though they note potential limitations related to fatty acid measurement compartments in the blood. Our finding also suggests caution in the use of pN3FA as a biomarker of dietary intake, since the dN3FA-pN3FA relationship that underlies this dietary proxy may be biased according to genetic variation.

Beyond this finding in the upstream analysis, we also report multiple genes passing a false discovery rate threshold in gene-level enrichment analysis of the downstream analysis linking pN3FA to hsCRP. Both *CBLL1* and *MICA* have biologically plausible explanations for impact on hsCRP via immune cell counts and function, establishing hypotheses for more in-depth exploration that would not have been prioritized using a standard GxE testing strategy without molecular mediators (all *p* > 0.05 in the standard pathway analysis). We also note that this downstream analysis has conceptual links to GxE studies using molecular quantities as exposures or “contexts” (Moore et al., 2019; Zhernakova et al., 2017).

In interpreting these results, it should be noted that the decomposition approach, including this N3FA application, depends on the quality and comprehensiveness of the measurement of mediating molecular species. For example, this study only considers N3FA as measured in plasma due to data availability, despite potential heterogeneity in genetic interaction results compare to other measurement compartments such as erythrocyte membranes (Smith et al., 2015). Additionally, despite the statistical significance of the uncovered *FADS* locus interaction, the practical relevance is quite small, with the interaction explaining only about 0.06% of the variance in pN3FA. Our use of only common variants in this study may have limited the space of possible variant effects; more clinically relevant effects may be discoverable by analyzing rare variation or using polygenic score-based approaches. Other approaches for future work include the expansion of this strategy to include multiple mediators, as has been described for omics-related main effect mediation analysis (Yang et al., 2021) but not extended to the realm of GxEs to our knowledge. Finally, the biological case study presented here explores a relatively narrow biological question involving the pathway from dN3FA to pN3FA to hsCRP; future analyses focused more directly on N3FA biology could include more fine-grained measurements of specific dietary and plasma N3FA subspecies as well as a broader range of outcome metrics.

In summary, our interaction decomposition approach leverages molecular mediators to improve the power to discover GxEs while improving the biological interpretability of the results. Applying this strategy using dietary and plasma N3FAs and hsCRP, we showed signal at the known *FADS* locus that was not present for the standard analysis, while additionally reporting multiple genes enriched in signal for the downstream analysis. We anticipate that this framework can guide more effective future GxE studies that leverage molecular quantities to increase discovery and mechanistic understanding.

## Data and code availability

Code supporting the simulations and analyses described here can be found at https://github.com/kwesterman/ukb-n3fa-decomp. The UK Biobank data can be obtained through application at https://www.ukbiobank.ac.uk/.

---

**Supplementary Figure S1:**
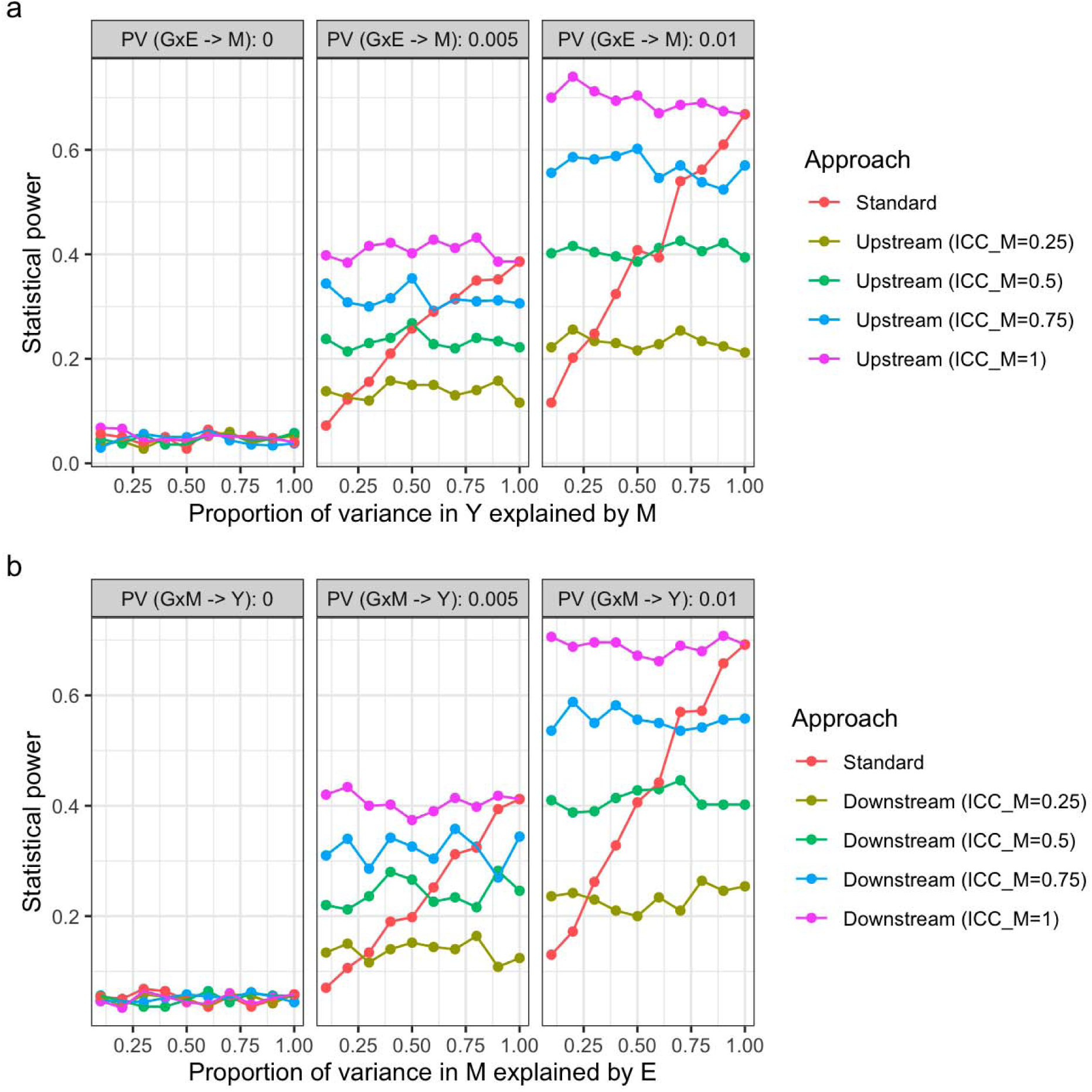
Statistical power simulation results for the decomposed versus standard approaches. *X*-axes correspond to the strength (quantified by proportion of variance explained) of the M-Y relationship (upstream, panel a) or E-M relationship (downstream, panel b). Faceted panels correspond to the strength of the simulated genetic interaction with E (upstream) or M (downstream). Colors correspond to the choice of test and associated measurement error in M (quantified by the ICC, where 0 indicates no signal and 1 indicates perfect measurement).

**Supplementary Figure S2:**
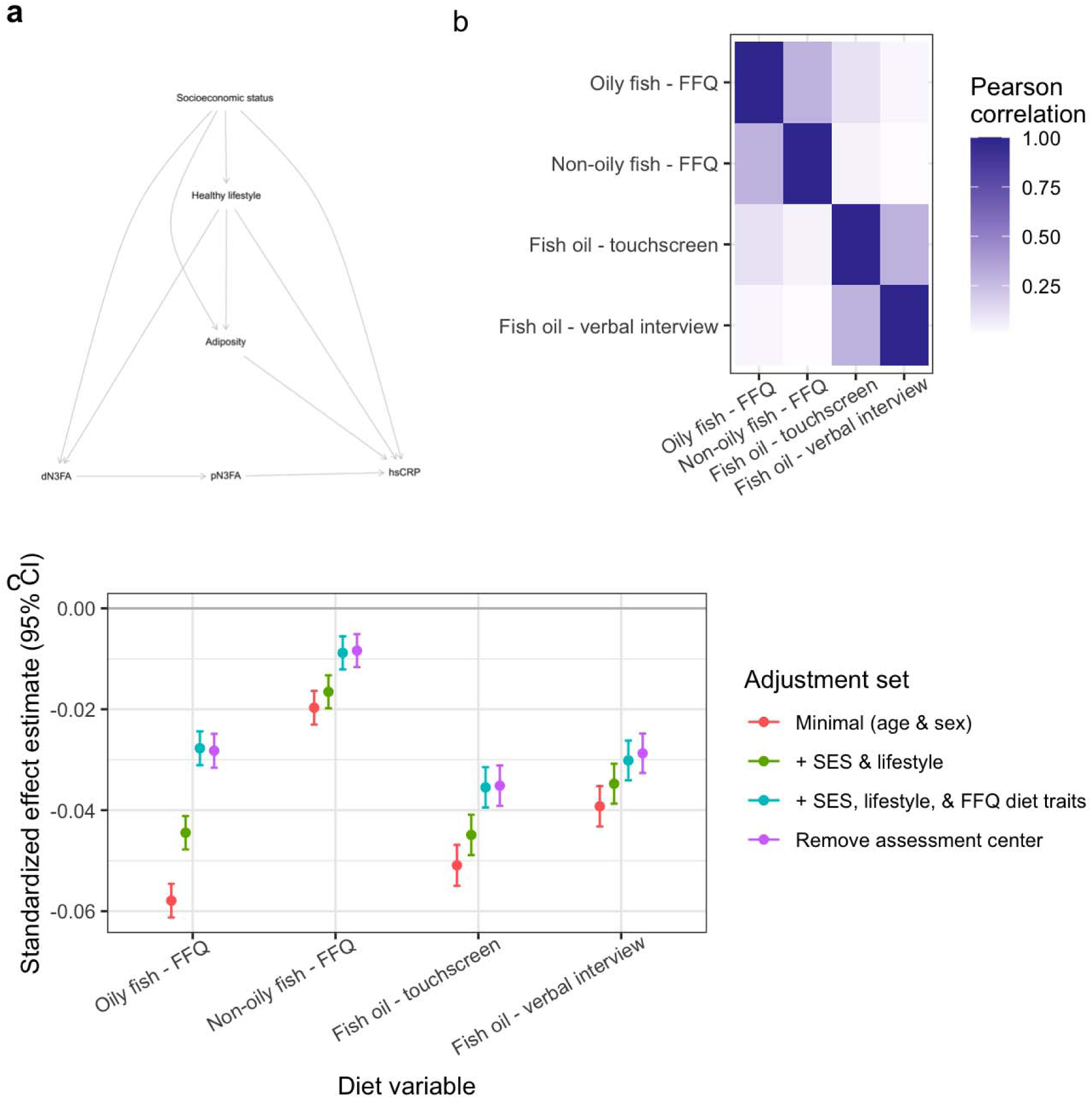
N3FA exposure and covariate selection. a) Directed acyclic graph informing covariate selection based on dN3FA-hsCRP main effect, with mediation by pN3FA. b) Heatmap of correlations between key N3FA-containing dietary variables (fish) and supplements (fish oil). c) Effect estimates for the relationship between these variables and log-transformed hsCRP (standardized to units of SD_outcome_/SD_exposure_). Colors correspond to confounder adjustment sets (SES = socioeconomic status indicators; purple points correspond to estimates adjusted for the full set of covariates minus all categorical indicators for assessment center; see Methods).

**Supplementary Figure S3:**
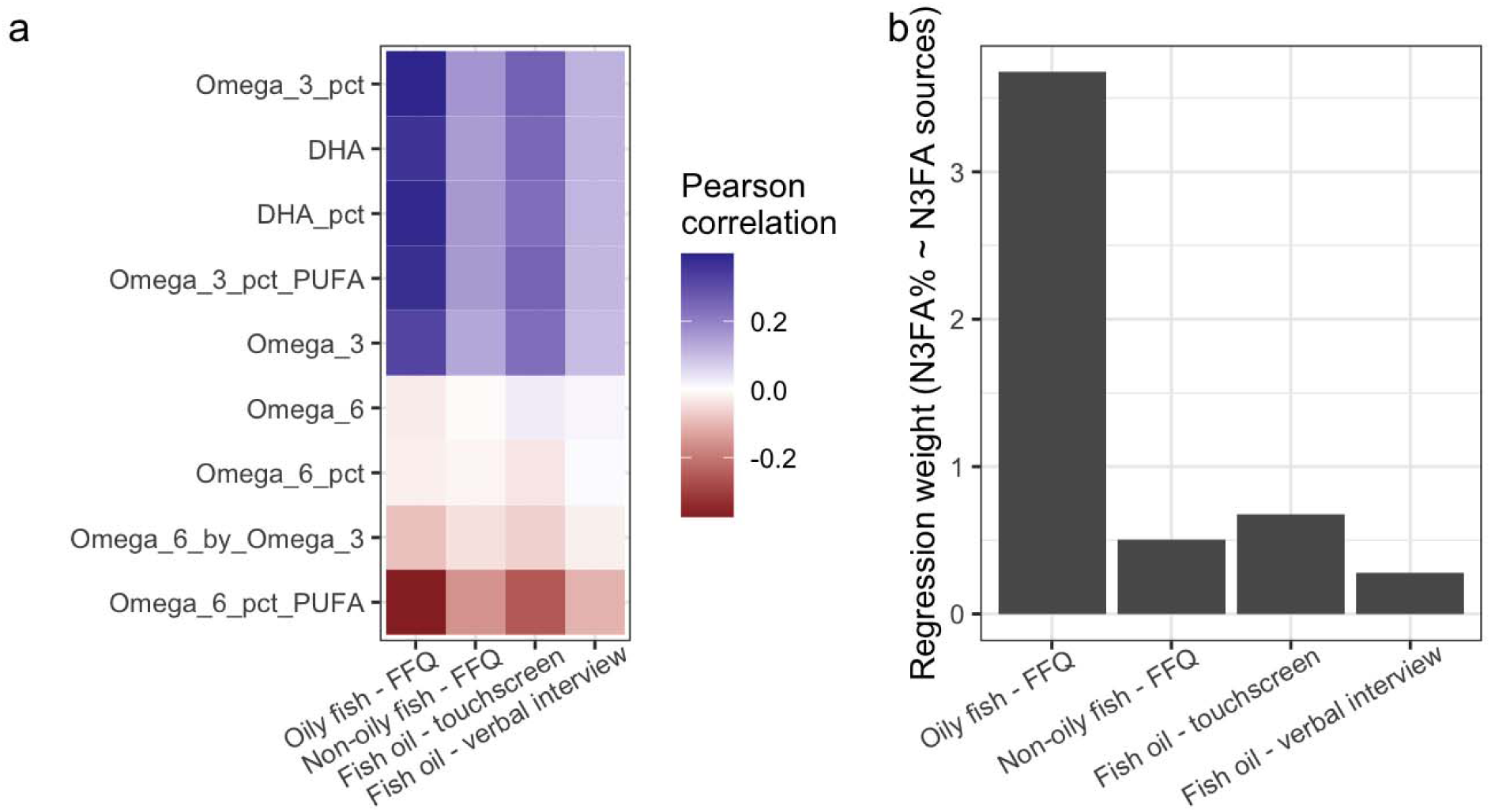
Mediator selection and imputation of dietary N3FA (dN3FA). a) Heatmap of correlations between fish and fish oil variables and various omega-3 and omega-6 species available in the Nightingale metabolomics data. b) Multivariable regression effect estimates for each of these dietary variables on pN3FA (plasma omega-3 as a percentage of total fatty acids). Units for exposures are servings/day for fish variables and binary yes/no indicators for fish oil variables.

**Supplementary Figure S4:**
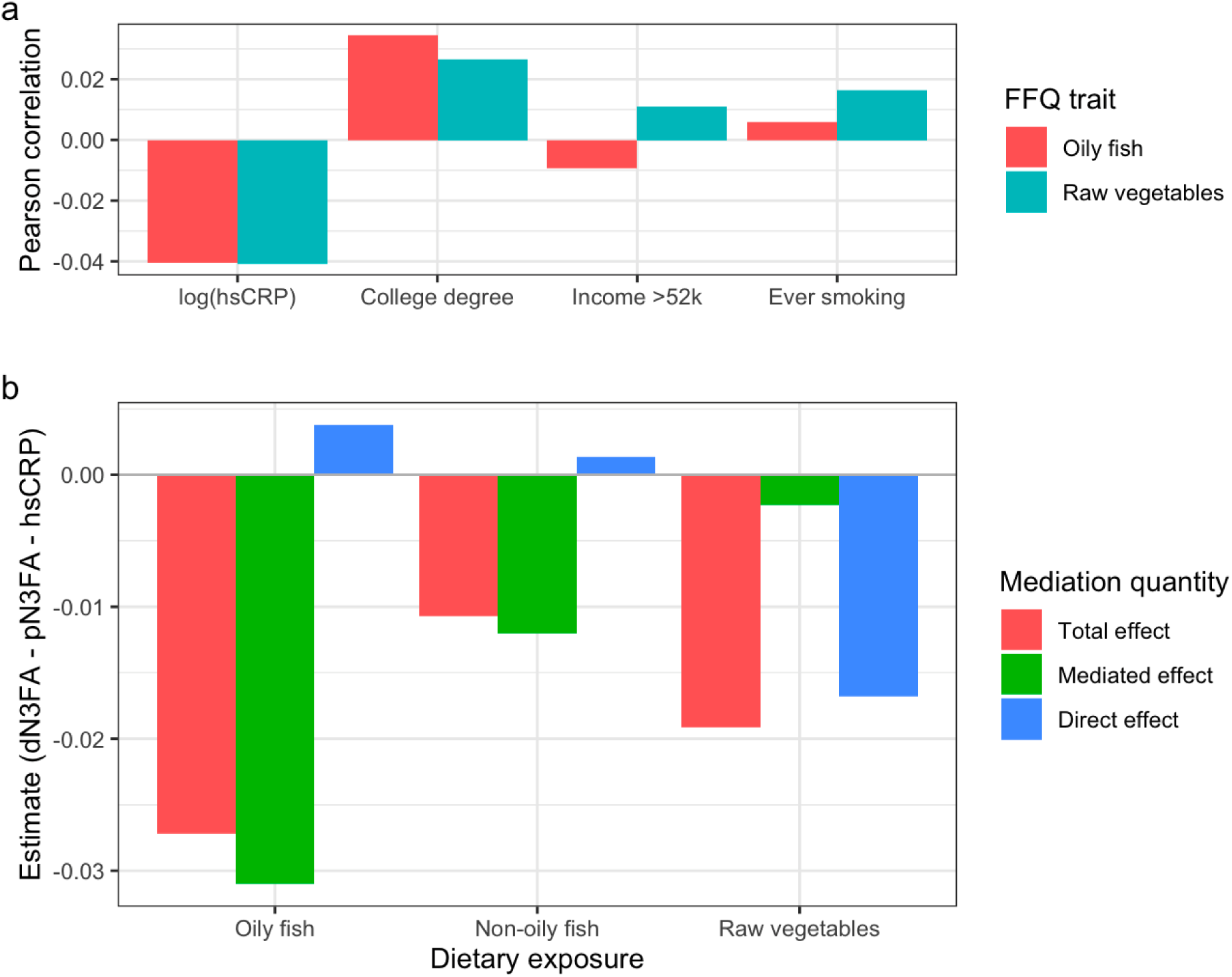
Raw vegetable intake as a negative control for mediation results. a) Barplot displays Pearson correlations between oily fish (variable of interest) and raw vegetable (negative control) intake and other variables of interest. b) Mediation testing results for the impact of oily fish (a primary N3FA source), non-oily fish (a modest N3FA source), and raw vegetable intake on log(hsCRP) through pN3FA as a mediator. Colors correspond to various estimates derived from mediation testing Monte Carlo draws (see Methods).

**Supplementary Figure S5:**
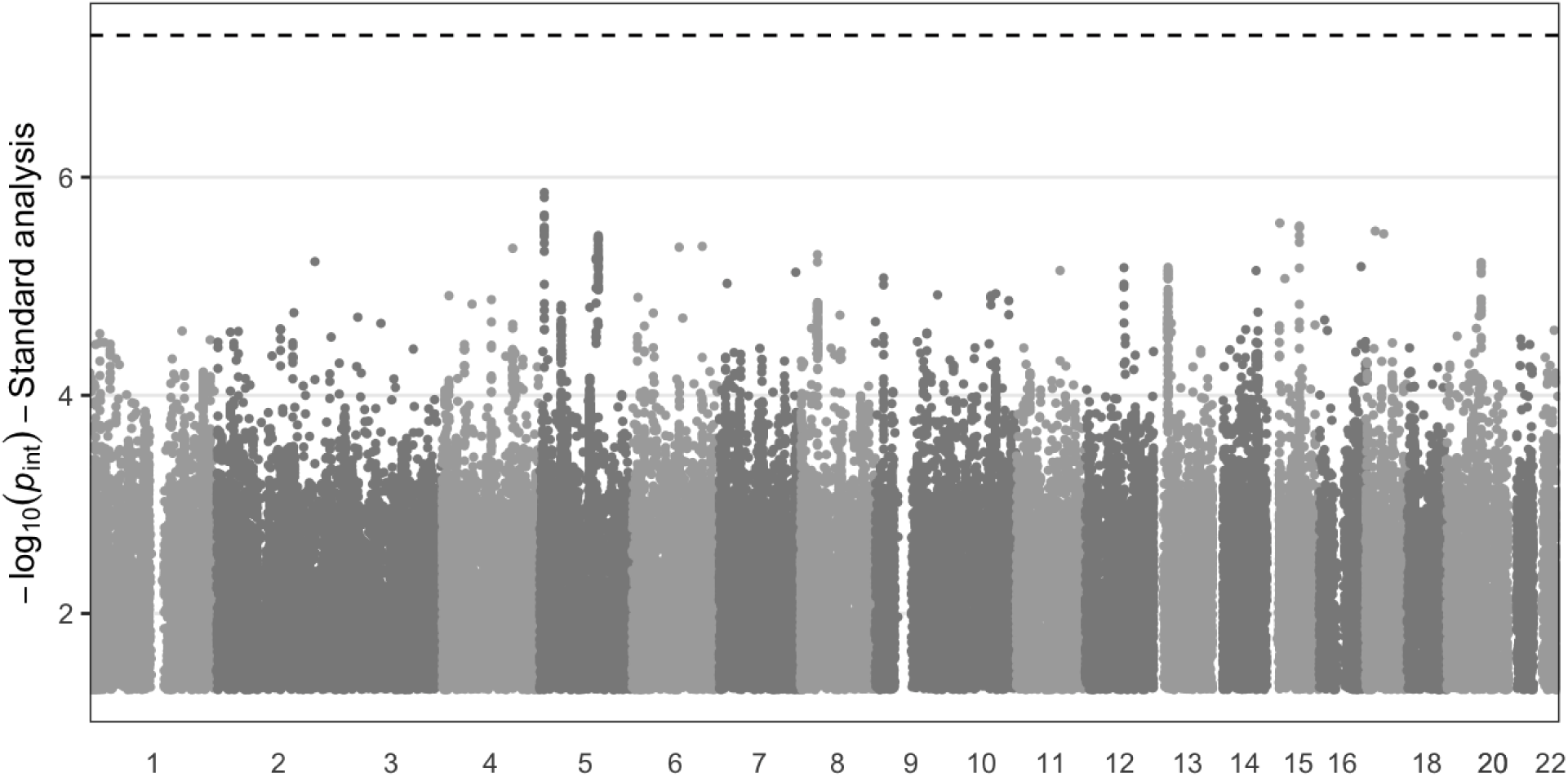
Manhattan plot displays variant interaction *p*-values as a function of chromosomal position for the standard analysis approach (dN3FA exposure and hsCRP outcome). The *y*-axis shows -log(*p*) for interaction tests (based on robust standard errors), while the *x*-axis indicates chromosomal position. Dotted line indicates the genome-wide significance threshold of 5×10^-8^.

## References

1. Abdelmagid, S. A., Clarke, S. E., Nielsen, D. E., Badawi, A., El-Sohemy, A., Mutch, D. M. & Ma, D. W. L. (2015). Comprehensive profiling of plasma fatty acid concentrations in young healthy canadian adults. PLoS ONE, 10(2). 10.1371/journal.pone.0116195

2. Baron, R. M. & Kenny, D. A. (1986). The moderator-mediator variable distinction in social psychological research: Conceptual, strategic, and statistical considerations. Journal of Personality and Social Psychology, 51(6). 10.1037//0022-3514.51.6.1173

3. Bradbury, K. E., Young, H. J., Guo, W. & Key, T. J. (2018). Dietary assessment in UK Biobank: an evaluation of the performance of the touchscreen dietary questionnaire. Journal of Nutritional Science, 7. 10.1017/jns.2017.66

4. Bycroft, C., Freeman, C., Petkova, D., Band, G., Elliott, L. T., Sharp, K., Motyer, A., Vukcevic, D., Delaneau, O., O’Connell, J., Cortes, A., Welsh, S., Young, A., Effingham, M., McVean, G., Leslie, S., Allen, N., Donnelly, P. & Marchini, J. (2018). The UK Biobank resource with deep phenotyping and genomic data. Nature, 562(7726), 203–209. 10.1038/s41586-018-0579-z

5. Calder, P. C. (2015). Marine omega-3 fatty acids and inflammatory processes: Effects, mechanisms and clinical relevance. In Biochimica et Biophysica Acta - Molecular and Cell Biology of Lipids (Vol. 1851, Issue 4). 10.1016/j.bbalip.2014.08.010

6. Chang, C. C., Chow, C. C., Tellier, L. C. A. M., Vattikuti, S., Purcell, S. M. & Lee, J. J. (2015). Second-generation PLINK: rising to the challenge of larger and richer datasets. GigaScience, 4(1), 7. 10.1186/s13742-015-0047-8

7. de Leeuw, C. A., Mooij, J. M., Heskes, T. & Posthuma, D. (2015). MAGMA: Generalized Gene-Set Analysis of GWAS Data. PLoS Computational Biology, 11(4). 10.1371/journal.pcbi.1004219

8. Gauderman, W. J., Mukherjee, B., Aschard, H., Hsu, L., Lewinger, J. P., Patel, C. J., Witte, J. S., Amos, C., Tai, C. G., Conti, D., Torgerson, D. G., Lee, S. & Chatterjee, N. (2017). Update on the State of the Science for Analytical Methods for Gene-Environment Interactions. American Journal of Epidemiology, 186(7), 762–770. 10.1093/aje/kwx228

9. Imai, K., Keele, L. & Tingley, D. (2010). A General Approach to Causal Mediation Analysis. Psychological Methods, 15(4). 10.1037/a0020761

10. Kasela, S., Aguet, F., Kim-Hellmuth, S., Brown, B. C., Nachun, D. C., Tracy, R. P., Durda, P., Liu, Y., Taylor, K. D., Johnson, W. C., Van Den Berg, D., Gabriel, S., Gupta, N., Smith, J. D., Blackwell, T. W., Rotter, J. I., Ardlie, K. G., Manichaikul, A., Rich, S. S., … Lappalainen, T. (2024). Interaction molecular QTL mapping discovers cellular and environmental modifiers of genetic regulatory effects. The American Journal of Human Genetics, 111(1), 133–149. 10.1016/j.ajhg.2023.11.013

11. Kidd, J., Raulerson, C. K., Mohlke, K. L. & Lin, D. Y. (2023). Mediation analysis of multiple mediators with incomplete omics data. Genetic Epidemiology, 47(1). 10.1002/gepi.22504

12. Kwan, J. L. Y. & Chan, W. (2018). Variable system: An alternative approach for the analysis of mediated moderation. Psychological Methods, 23(2). 10.1037/met0000160

13. Lemaitre, R. N., Tanaka, T., Tang, W., Manichaikul, A., Foy, M., Kabagambe, E. K., Nettleton, J. A., King, I. B., Weng, L. C., Bhattacharya, S., Bandinelli, S., Bis, J. C., Rich, S. S., Jacobs, D. R., Cherubini, A., McKnight, B., Liang, S., Gu, X., Rice, K., … Steffen, L. M. (2011). Genetic loci associated with plasma phospholipid N-3 fatty acids: A Meta-Analysis of Genome-Wide association studies from the charge consortium. PLoS Genetics, 7(7). 10.1371/journal.pgen.1002193

14. Little, T. D., Card, N. A., Bovaird, J. A., Preacher, K. J. & Crandall, C. S. (2012). Structural equation modeling of mediation and moderation with contextual factors. In Modeling Contextual Effects in Longitudinal Studies. 10.4324/9780203936825

15. MacKinnon, D. P., Krull, J. L. & Lockwood, C. M. (2000). Equivalence of the mediation, confounding and suppression effect. Prevention Science, 1(4). 10.1023/A:1026595011371

16. Mohammadi-Shemirani, P., Sood, T. & Paré, G. (2023). From ‘Omics to Multi-omics Technologies: the Discovery of Novel Causal Mediators. In Current Atherosclerosis Reports (Vol. 25, Issue 2). 10.1007/s11883-022-01078-8

17. Moore, R., Casale, F. P., Jan Bonder, M., Horta, D., Franke, L., Barroso, I. & Stegle, O. (2019). A linear mixed-model approach to study multivariate gene–environment interactions. Nature Genetics, 51(1), 180–186. 10.1038/s41588-018-0271-0

18. Patel, C. J., Kerr, J., Thomas, D. C., Mukherjee, B., Ritz, B., Chatterjee, N., Jankowska, M., Madan, J., Karagas, M. R., Mcallister, K. A., Mechanic, L. E., Fallin, M. D., Ladd-Acosta, C., Blair, I. A., Teitelbaum, S. L. & Amos, C. I. (2017). Opportunities and challenges for environmental exposure assessment in population-based studies. In Cancer Epidemiology Biomarkers and Prevention (Vol. 26, Issue 9). 10.1158/1055-9965.EPI-17-0459

19. R Core Team. (2022). R: A Language and Environment for Statistical Computing (4.2.2). https://www.R-project.org/

20. Ritchie, S. C., Surendran, P., Karthikeyan, S., Lambert, S. A., Bolton, T., Pennells, L., Danesh, J., Di Angelantonio, E., Butterworth, A. S. & Inouye, M. (2023). Quality control and removal of technical variation of NMR metabolic biomarker data in ∼120,000 UK Biobank participants. Scientific Data, 10(1). 10.1038/s41597-023-01949-y

21. Schulze, M. B., Minihane, A. M., Saleh, R. N. M. & Risérus, U. (2020). Intake and metabolism of omega-3 and omega-6 polyunsaturated fatty acids: nutritional implications for cardiometabolic diseases. In The Lancet Diabetes and Endocrinology (Vol. 8, Issue 11). 10.1016/S2213-8587(20)30148-0

22. Smith, C. E., Follis, J. L., Nettleton, J. A., Foy, M., Wu, J. H. Y., Ma, Y., Tanaka, T., Manichakul, A. W., Wu, H., Chu, A. Y., Steffen, L. M., Fornage, M., Mozaffarian, D., Kabagambe, E. K., Ferruci, L., Chen, Y. D. I., Rich, S. S., Djoussé, L., Ridker, P. M., … Lemaitre, R. N. (2015). Dietary fatty acids modulate associations between genetic variants and circulating fatty acids in plasma and erythrocyte membranes: Meta-analysis of nine studies in the CHARGE consortium. Molecular Nutrition and Food Research, 59(7). 10.1002/mnfr.201400734

23. Sudlow, C., Gallacher, J., Allen, N., Beral, V., Burton, P., Danesh, J., Downey, P., Elliott, P., Green, J., Landray, M., Liu, B., Matthews, P., Ong, G., Pell, J., Silman, A., Young, A., Sprosen, T., Peakman, T. & Collins, R. (2015). UK Biobank: An Open Access Resource for Identifying the Causes of a Wide Range of Complex Diseases of Middle and Old Age. PLOS Medicine, 12(3), e1001779. 10.1371/journal.pmed.1001779

24. Tingley, D., Yamamoto, T., Hirose, K., Keele, L. & Imai, K. (2014). Mediation: R package for causal mediation analysis. Journal of Statistical Software, 59(5). 10.18637/jss.v059.i05

25. Westerman, K. E., Pham, D. T., Hong, L., Chen, Y., Sevilla-González, M., Sung, Y. J., Sun, Y. V, Morrison, A. C., Chen, H. & Manning, A. K. (2021). GEM: scalable and flexible gene–environment interaction analysis in millions of samples. Bioinformatics. 10.1093/bioinformatics/btab223

26. Yang, T., Niu, J., Chen, H. & Wei, P. (2021). Estimation of total mediation effect for high-dimensional omics mediators. BMC Bioinformatics, 22(1). 10.1186/s12859-021-04322-1

27. Zhernakova, D. V., Deelen, P., Vermaat, M., Van Iterson, M., Van Galen, M., Arindrarto, W., Van’t Hof, P., Mei, H., Van Dijk, F., Westra, H. J., Bonder, M. J., Van Rooij, J., Verkerk, M., Jhamai, P. M., Moed, M., Kielbasa, S. M., Bot, J., Nooren, I., Pool, R., … Franke, L. (2017). Identification of context-dependent expression quantitative trait loci in whole blood. Nature Genetics, 49(1). 10.1038/ng.3737

